# A Polygenic Score for Reduced Kidney Function and Adverse Outcomes in a Chronic Kidney Disease Cohort

**DOI:** 10.1101/2022.06.02.22275913

**Authors:** Inga Steinbrenner, Zhi Yu, Jin Jin, Ulla T. Schultheiss, Fruzsina Kotsis, Morgan E. Grams, Josef Coresh, Matthias Wuttke, GCKD investigators, Florian Kronenberg, Kai-Uwe Eckardt, Nilanjan Chatterjee, Peggy Sekula, Anna Köttgen

## Abstract

We studied whether a polygenic score for reduced kidney function developed from population-based studies was associated with adverse outcomes among persons with chronic kidney disease. The polygenic score was significantly associated with incident kidney failure, major adverse cardiovascular outcomes and overall mortality while adjusting for age, sex, and baseline eGFR: the hazard ratio for kidney failure over 6.5 years was 1.83 (95% CI 1.40-2.39) comparing those in the highest and lowest quartiles of the polygenic score. However, the significant associations of the polygenic score did not translate to improved outcome prediction in comparison to established risk equations.

## Introduction

Chronic kidney disease (CKD) is a global health burden of increasing importance that affects >10% of the general adult population.^1^ CKD is defined as the sustained presence of abnormalities of kidney structure or function, and classified using estimated glomerular filtration rate (eGFR) and the urinary albumin-to-creatinine ratio (UACR).^2^ The eGFR based on serum creatinine is the most common measure of kidney function. Persons with CKD are at increased risk of adverse outcomes such as kidney failure (KF), cardiovascular diseases, and premature death.^3^

Genome-wide association studies enable the calculation of polygenic scores (PGS). PGS aggregate the effects of many genetic variants into a single number and permit a straight-forward investigation of the association between a genetic predisposition with a given outcome. Yet, evaluations of such PGS in external studies, including different target populations, are often limited. Large CKD cohorts like the prospective German Chronic Kidney Disease (GCKD)^4^ study represent a valuable opportunity to study whether a PGS for reduced eGFR (termed “eGFR PGS”), is associated with adverse outcomes in CKD. Therefore, the aim of this study was to investigate if (i) a polygenic predisposition to lower eGFR is associated with KF, cardiovascular events and mortality in persons with CKD, and if (ii) the eGFR PGS carries predictive ability by itself and in addition to established risk factors.

## Short Methods

We developed and tuned a PGS for log(eGFR) in independent, general population-based study samples consisting of European ancestry participants of the UK Biobank^5^ and the CKDGen Consortium^6^ via the LDpred algorithm^7^ as described in the **Supplementary Methods**, following a workflow published previously^8^. The best performing PGS was then applied using data from 4 924 GCKD participants with moderate CKD and full information. The eGFR PGS was calculated, standardised and evaluated for association with i) KF defined as a composite endpoint of kidney replacement therapy (dialysis, kidney transplantation) or death due to foregoing dialysis, ii) a composite endpoint of acute myocardial infarction, cerebral haemorrhage and stroke (short: 3P-MACE), and iii) overall mortality (short: death). Secondary outcomes included the three components of 3P-MACE. Cox proportional hazard regression was conducted to estimate (cause-specific) hazard ratios. In case of KF and 3P-MACE and its components, we considered death of other causes as a competing event and additionally analysed the subdistribution hazard for the event of interest. For each outcome, we adjusted for three different sets of variables: 1) age + sex + significant genetic principal component, 2) model 1 + baseline eGFR and 3) model 2 + log(UACR). Predictive performance of the eGFR PGS for KF, above and beyond the well-established kidney failure risk equation (KFRE; based on age, sex, eGFR and UACR) introduced by Tangri *et al*.^9^, was assessed using prediction error curves, the c-index, a calibration plot and the 6-year receiver operating characteristic (ROC) curve.^10,11^ The KFRE was chosen for comparison because of its common use in the care for CKD patients. The statistical significance level was Bonferroni-corrected for three adjustments (p<0.05/3). If not stated otherwise, results of model 2) are reported. Detailed methods can be found in the **Supplementary Methods** section of the **Supplementary Materials**.

## Results

In the PGS development stage, the PGS with the best performance (**Supplementary Methods**) assumed a fraction of causal variants of 0.1 and was subsequently applied in the GCKD study. The mean eGFR among 4 924 participants (39.8% women) with complete data was 49.4 mL/min/1.73m^2^ (**Supplementary Table 1**). As expected in a CKD cohort that recruited mostly participants with stage G3 CKD, Spearman correlation of the eGFR PGS with eGFR at the enrolment visit was low (0.03). Over a median follow-up of 6.5 years (Q1: 6.5; Q3: 6.5), 470 (9.5%) participants experienced KF, 551 (11.2%) 3P-MACE, and 630 (12.8%) died.

### A PGS for eGFR is associated with adverse outcomes

A higher continuous PGS, corresponding to a lifelong genetic predisposition to lower eGFR, was associated with higher risk for all three outcomes (KF: HR 1.22, 95% confidence interval (CI) [1.11;1.33], 3P-MACE: 1.15 [1.06;1.25], death: 1.13 [1.04;1.22]) after controlling for age, sex, baseline eGFR and one signifcant genetic principal component (model 2; **Figure** 1, panel A; **Supplementary Tables 2-4**). Upon adjustment for the UACR (model 3), these associations remained significant for KF and 3P-MACE, but were attenuated for death (KF: HR 1.12, 95% CI [1.02;1.23], 3P-MACE: 1.12 [1.03;1.22], death: 1.09 [1.01;1.18]). Results without adjustment for eGFR (model 1) were similar to model 2. The subdistribution analyses delivered similar results, indicating the absence of indirect effects through the competing event. The risk of KF over 6.5 years was 1.83 (95% CI 1.40-2.39) times higher comparing those in the highest and lowest PGS quartiles (**Supplementary Table 2**). Across deciles of the eGFR PGS, participants in the highest decile, corresponding to genetically lowest eGFR, had a more than two-fold increased risk of KF compared to those in the lowest decile (HR=2.17 [1.41;3.33]).

The unadjusted cumulative incidence function for KF as well as for 3P-MACE was most pronounced for the highest quartile of the eGFR PGS, in line with the Cox regression results (**Figure 1**, panels B and C). Notably, the functions clearly distinguished participants in the highest and lowest quartiles of the PGS (cumulative incidence of KF of 13.8% (highest quartile) vs. 7.7% (lowest quartile) and 14.6% vs 10.9% for 3P-MACE). The same trends were observed for the 3P-MACE sub-outcomes acute myocardial infarction, cerebral haemorrhage and stroke (**Supplementary Table 4**), albeit mostly not significant.

**Figure 1.**
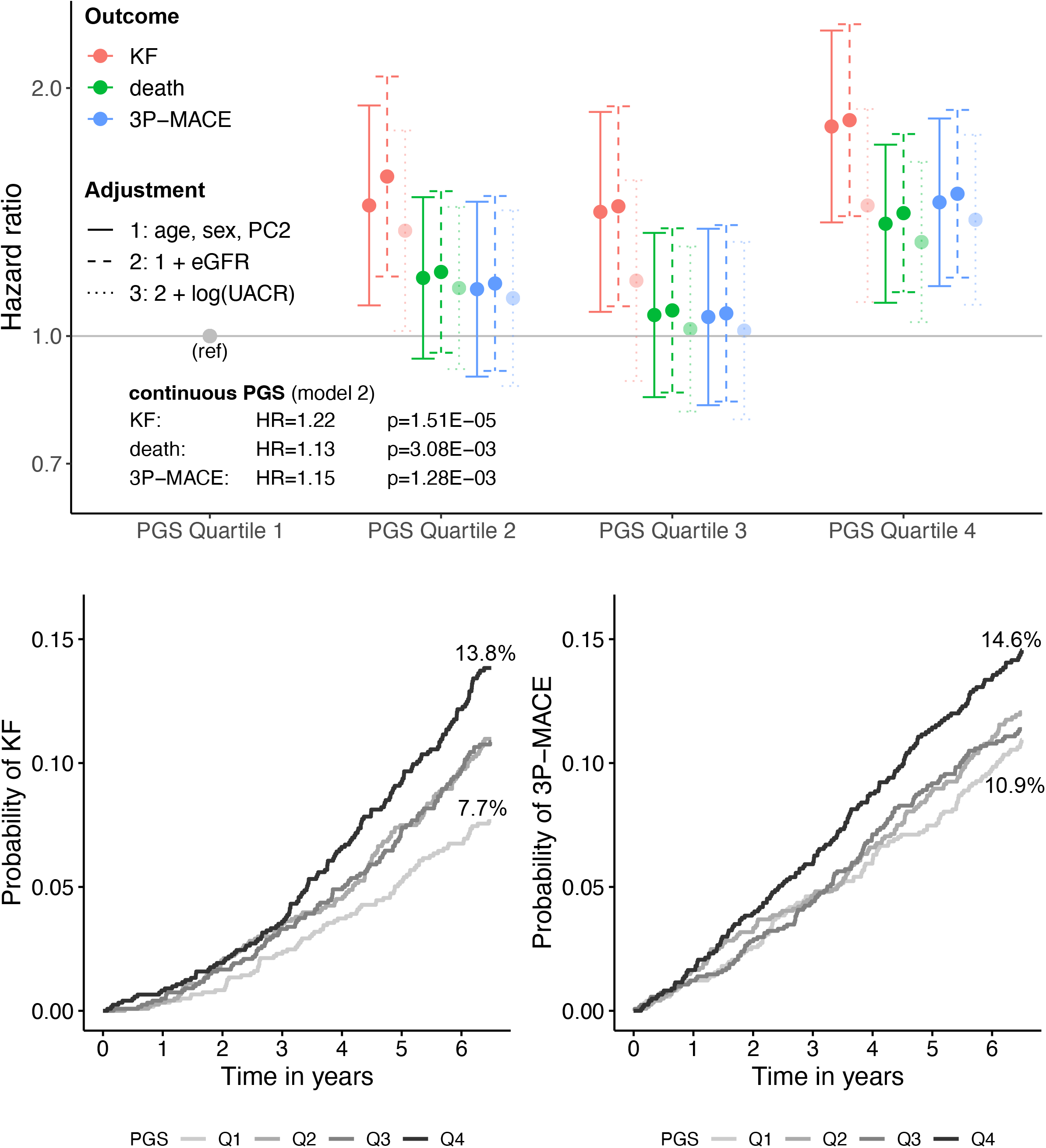
Association results of the eGFR PGS with all studied outcomes. **A**: Hazard ratios by PGS quartiles for different adjustment models. The studied outcomes are color-coded. **B**: Cumulative incidence function (unadjusted) for KF by quartile of PGS. **C**: Cumulative incidence function (unadjusted) for 3P-MACE by quartile of PGS.

### Predictive ability of the PGS

The likelihood ratio test (LRT) comparing the well-established KFRE for incident KF with the same model after addition of the PGS was nominally significant (**Supplementary Table 5**), indicating potential for improvement of model performance. Yet, neither calibration nor the prediction error curves, the c-index or the receiver operating characteristic (ROC) curve at year 6 improved when adding the PGS to the KFRE model for KF (**Figure 2**). Addition of the PGS to the KFRE resulted in no change of the c-index of 0.84 for KF. Similarly, the integrated prediction error curve (IPEC), where higher values indicate worse prediction, changed from 56.37 to 56.72 for KF.

**Figure 2.**
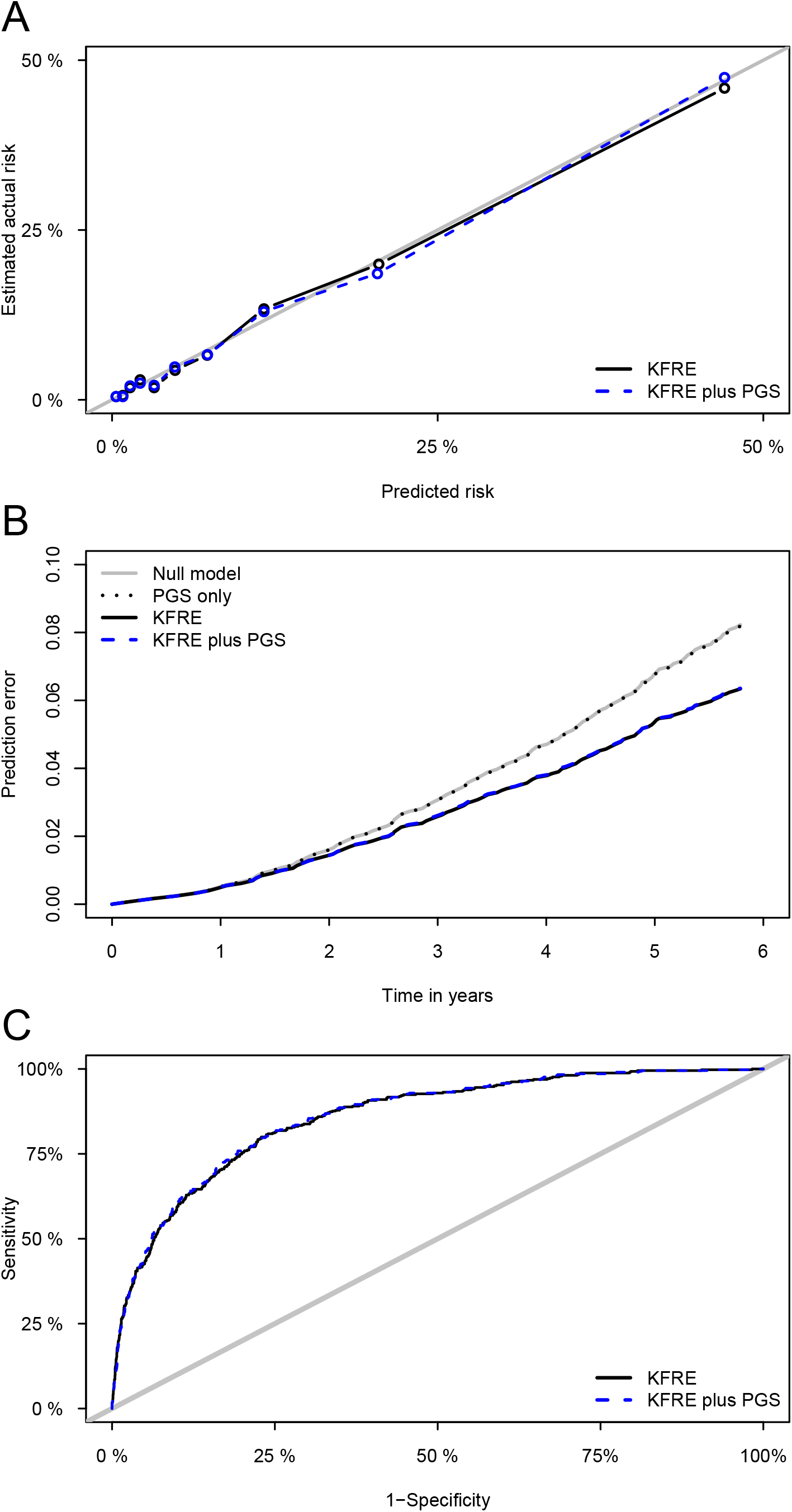
Predictive ability of the eGFR PGS. **A**: Calibration plot at year six; **B**: Prediction error curves for KF over time; **C**: Six-year ROC curve for KF; Each panel displays different adjustments: Null model: unadjusted; PGS only: PGS; KFRE: adjustment for age, sex, baseline eGFR, log(UACR); KFRE plus PGS: KFRE + PGS.

## Discussion

In this study of an eGFR PGS in persons with CKD, we found significant associations with KF, 3P-MACE and death but limited predictive ability with respect to KF.

While various studies focused primarily on the association of an eGFR PGS with clinical outcomes or the prediction of the presence of CKD (e.g. Yun *et al*.^12^, Khan *et al*.^13^) in the general population, little is known about the relationship of an eGFR PGS to adverse outcomes in persons with already diagnosed moderate CKD, a novel aspect addressed in this study. In agreement with our findings, a previous population-based study by Yu *et al*.^8^ reported significant positive association of a similarly developed eGFR PGS with incident CKD and KF. Thus, our results support that an eGFR PGS developed in the general population is associated with adverse kidney outcomes not only in population-based studies, but also among persons with already existing CKD.

At the same time, this study showed no improvement in risk prediction, underlining the finding of Gleiss *et al*.^14^ that significant associations are only a necessary but not a sufficient condition for the usefulness of prognostic markers in prediction studies. The limited use of PGS for risk prediction has been pointed out by Wald *et al*. previously.^15^ While several studies concluded that eGFR PGS were suitable for prediction of the presence of CKD^8,13^, they did not evaluate prediction of incident outcomes, although these are two important separate aspects in PGS validity according to Torkamani *et al*.^16^ A previous population-based prospective study of incident CKD^8^ on the other hand did not report on risk prediction. Thus, evidence around the predictive ability of an eGFR PGS for adverse outcomes among CKD patients is lacking, a knowledge gap filled by our study. Our results suggests that in order to achieve the goal of clinical translation of an eGFR PGS^17^, further validation in CKD cohorts is warranted in order to identify settings in which such a PGS can add above and beyond established risk prediction equations that do not require genetic information.

Strengths of our study are its new focus on persons with CKD and a large sample size with systematic outcome ascertainment over 6.5 years of follow-up. Despite the large number of outcomes, however, the power to study finer distinctions, such as deciles of the PGS or rare outcomes, was limited, as discussed previously.^18^ Another limitation is that findings in the GCKD study may not be generalizable to non-European ancestry populations.

In conclusion, our study revealed significant associations between a polygenic predisposition to lower eGFR and KF, 3P-MACE, and death among persons with moderate CKD, emphasising the importance of genetic background even after disease onset. However, the eGFR PGS carried no added predictive ability with regard to KF beyond the well-performing KFRE among patients with established CKD.

## Data Availability

Public posting of individual level participant data is not covered by the informed patient consent form. As stated in the patient consent form and approved by the Ethics Committees, a dataset containing pseudonyms can be obtained
by collaborating scientists upon approval of a scientific project proposal by the steering committee of the GCKD study: https//:www.gckd.org. All data produced in the present study are available upon reasonable request to the authors.

## Acknowledgements and funding

The GCKD study was funded by grants from the BMBF (grant number 01ER0804) and the KfH Foundation for Preventive Medicine and corporate sponsors (http://www.gckd.org). Genotyping in GCKD was supported by Bayer Pharma AG. We are grateful for the willingness of the patients to participate in the GCKD study. The enormous effort of the study personnel of the various regional centers is highly appreciated. We thank the large number of nephrologists who provide routine care for the patients and collaborate with the GCKD study. The GCKD Investigators are listed in the **Supplementary Material**.

## References

1. Eckardt KU, Coresh J, Devuyst O, et al. Evolving importance of kidney disease: from subspecialty to global health burden. Lancet Lond Engl. 2013;382(9887):158–169. doi:10.1016/S0140-6736(13)60439-0

2. Kidney Disease: Improving Global Outcomes (KDIGO) CKD Work Group. KDIGO 2012 Clinical Practice Guideline for the Evaluation and Management of Chronic Kidney Disease. Kidney inter. 2013;3:1–150.

3. Jha V, Garcia-Garcia G, Iseki K, et al. Chronic kidney disease: global dimension and perspectives. The Lancet. 2013;382(9888):260–272. doi:10.1016/S0140-6736(13)60687-X

4. Eckardt KU, Bärthlein B, Baid-Agrawal S, et al. The German Chronic Kidney Disease (GCKD) study: design and methods. Nephrol Dial Transplant Off Publ Eur Dial Transpl Assoc - Eur Ren Assoc. 2012;27(4):1454–1460. doi:10.1093/ndt/gfr456

5. Bycroft C, Freeman C, Petkova D, et al. The UK Biobank resource with deep phenotyping and genomic data. Nature. 2018;562(7726):203–209. doi:10.1038/s41586-018-0579-z

6. Köttgen A, Pattaro C. The CKDGen Consortium: ten years of insights into the genetic basis of kidney function. Kidney Int. 2020;97(2):236–242. doi:10.1016/j.kint.2019.10.027

7. Vilhjálmsson BJ, Yang J, Finucane HK, et al. Modeling Linkage Disequilibrium Increases Accuracy of Polygenic Risk Scores. Am J Hum Genet. 2015;97(4):576–592. doi:10.1016/j.ajhg.2015.09.001

8. Yu Z, Jin J, Tin A, et al. Polygenic Risk Scores for Kidney Function and Their Associations with Circulating Proteome, and Incident Kidney Diseases. J Am Soc Nephrol JASN. Published online September 21, 2021:ASN.2020111599. doi:10.1681/ASN.2020111599

9. Tangri N. A Predictive Model for Progression of Chronic Kidney Disease to Kidney Failure. JAMA. 2011;305(15):1553. doi:10.1001/jama.2011.451

10. Binder H, Schumacher M. Adapting prediction error estimates for biased complexity selection in high-dimensional bootstrap samples. Stat Appl Genet Mol Biol. 2008;7(1):Article12. doi:10.2202/1544-6115.1346

11. Gerds TA, Kattan MW, Schumacher M, Yu C. Estimating a time-dependent concordance index for survival prediction models with covariate dependent censoring. Stat Med. 2013;32(13):2173–2184. doi:10.1002/sim.5681

12. Yun S, Han M, Kim HJ, et al. Genetic risk score raises the risk of incidence of chronic kidney disease in Korean general population-based cohort. Clin Exp Nephrol. 2019;23(8):995–1003. doi:10.1007/s10157-019-01731-8

13. Khan A, Turchin MC, Patki A, et al. Genome-Wide Polygenic Score with APOL1 Risk Genotypes Predicts Chronic Kidney Disease across Major Continental Ancestries. Genetic and Genomic Medicine; 2021. doi:10.1101/2021.10.25.21265398

14. Gleiss A, Zeillinger R, Braicu EI, Trillsch F, Vergote I, Schemper M. Statistical controversies in clinical research: the importance of importance. Ann Oncol. 2016;27(7):1185–1189. doi:10.1093/annonc/mdw159

15. Wald NJ, Old R. The illusion of polygenic disease risk prediction. Genet Med. 2019;21(8):1705–1707. doi:10.1038/s41436-018-0418-5

16. Torkamani A, Wineinger NE, Topol EJ. The personal and clinical utility of polygenic risk scores. Nat Rev Genet. 2018;19(9):581–590. doi:10.1038/s41576-018-0018-x

17. Hao L, Kraft P, Berriz GF, et al. Development of a clinical polygenic risk score assay and reporting workflow. Nat Med. 2022;28(5):1006–1013. doi:10.1038/s41591-022-01767-6

18. Liu L, Kiryluk K. Genome-wide polygenic risk predictors for kidney disease. Nat Rev Nephrol. 2018;14(12):723–724. doi:10.1038/s41581-018-0067-6

