## Supplemental Material for "A Polygenic Score for Reduced Kidney Function and Adverse Outcomes in a Chronic Kidney Disease Cohort"

### Supplementary Material

**Supplementary Tables:** See separate Excel File.

**Supplementary Note: List of GCKD Participating Institutions and Investigators.**

The nine GCKD participating institutions are: RWTH Aachen University, Aachen, Charité – University-Medicine, Berlin, Friedrich-Alexander University, Erlangen, Albert-Ludwigs-University, Freiburg, Friedrich-Schiller University, Jena, Hannover Medical School, Hannover, Medical Faculty, Ruprecht-Karls University, Heidelberg, Medical Faculty, Ludwig-Maximilians-University, Munich, and Julius-Maximilians-University, Würzburg.

A list of nephrologists currently collaborating with the GCKD study is available at www.gckd.org.

- University of Erlangen-Nürnberg
  Kai-Uwe Eckardt, Heike Meiselbach, Markus P. Schneider, Mario Schiffer, Hans-Ulrich Prokosch, Barbara Bärthlein, Andreas Beck, André Reis, Arif B. Ekici, Susanne Becker, Ulrike Alberth-Schmidt, Anke Weigel; Sabine Marschall, Eugenia Schefler

- University of Freiburg
  Gerd Walz, Anna Köttgen, Ulla T. Schultheiß, Fruzsina Kotsis, Simone Meder, Erna Mitsch, Ursula Reinhard

- RWTH Aachen University
  Jürgen Floege, Turgay Saritas

- Charité, University Medicine Berlin
  Elke Schaeffner, Seema Baid-Agrawal, Kerstin Theisen

- Hannover Medical School
  Hermann Haller

- University of Heidelberg
  Martin Zeier, Claudia Sommerer, Johanna Theilinger

- University of Jena
  Gunter Wolf, Martin Busch, Rainer Paul

- Ludwig-Maximilians University of München
  Thomas Sitter

- University of Würzburg
  Christoph Wanner, Vera Krane, Antje Börner-Klein, Britta Bauer

- Medical University of Innsbruck, Division of Genetic Epidemiology
  Florian Kronenberg, Julia Raschenberger, Barbara Kollerits, Lukas Forer, Sebastian Schönherr, Hansi Weissensteiner

- University of Regensburg, Institute of Functional Genomics
  Peter Oefner, Wolfram Gronwald

- Institute of Medical Biometry, Informatics and Epidemiology, Medical Faculty, University of Bonn:
  Matthias Schmid, Jennifer Nadal

### Supplementary Methods

#### Supplementary Methods 1: Study design and participants

From 2010 to 2012, 5 217 participants with moderate CKD were enrolled in the prospective observational German Chronic Kidney Disease (GCKD) study.[1] A detailed description of the study population and design has been published.[2] Patients under regular care by a nephrologist were enrolled if they had 1) an eGFR between 30–60 mL/min per 1.73 $m^2$ or 2) an eGFR >60 mL/min per 1.73 $m^2$ in combination with albuminuria (urinary albumin/creatinine >300 mg/g or albuminuria >300 mg/day) or proteinuria (urinary protein/creatinine >500 mg/g or proteinuria >500 mg/day). Clinical endpoint collection is still ongoing, recorded continuously and adjudicated in a standardised fashion. The GCKD Study was registered in the national registry for clinical studies (DRKS 00003971) and approved by all local ethic committees.

Outcomes of interest in this study were kidney failure (KF), defined as a combined outcome of dialysis, kidney transplantation and death due to forgoing dialysis, a combined outcome of acute myocardial infarction, cerebral haemorrhage and stroke (short: 3P-MACE), as well as overall mortality (short: death). Further, myocardial infarction, cerebral haemorrhage and stroke were analysed separately as secondary outcomes. Data on covariates and outcomes over the first six years of follow-up were available in this project for 4 873 study participants.

The UK Biobank (UKBB) is a prospective study involving around half a million adult people aged 40-69 at enrollment in the UK, with anonymised data made publicly available for approved researchers.[3] The comprehensive data base includes health record data, blood and saliva samples, life-style data as well as genetic data.

The CKDGen Consortium is an international effort of researchers working on epidemiological studies with genome-wide genetic data and kidney function measurements.[4] Their goal is to gain insights into the genetic mechanisms underlying kidney function and disease.

**Supplementary Methods 2: Definition of baseline variables in GCKD**

In short, variables evaluated at baseline were defined as follows (methods and definitions were previously published[1,2]):

- Serum creatinine was measured using an IDMS traceable enzymatic assay (Creatinine Plus, Roche).
- The Chronic Kidney Disease Epidemiology Collaboration (CKD-EPI) formula[5] was used to estimate GFR.
- The urinary albumin-to-creatinine ratio (UACR) was calculated from urinary creatinine (IDMS traceable enzymatic assay [Creatinine Plus, Roche]) and albumin (ALBU-XS assay [Roche/Hitachi Diagnostics GmbH, Mannheim, Germany]).
- High-sensitivity C-reactive protein (CRP) was measured using an immunoturbidimetric test (CRPHS, Roche, Germany) on a Roche/Hitachi MODULAR (P).
- Total and high-density lipoprotein cholesterol (HDL) as well as triglycerides were measured using an enzymatic colorimetric method (CHOD-PAP, Roche, Germany) on a Roche/Hitachi MODULAR (P).
- Patients were considered as smokers if they reported current daily or occasional smoking.
- Blood pressure medication was based on reported medication use (Anatomical Therapeutic Chemical [ATC] codes beginning with either of the following: ‘C02’, ‘C03’, ‘C07’, ‘C08’, ‘C09’).
- Systolic blood pressure was calculated as the mean out of three measurements after five minutes resting using a standardised device (OMRON M5 Professional, Mannheim, Germany).

- Diabetes was based on a baseline serum hemoglobin A1c (HbA1c) of ≥ 6.5% or self-documented intake of diabetes medication (ATC codes beginning with ‘A10’).
- History of cardiovascular disease was defined as a positive history of any of the following: stroke, carotid artery operation/stenting, peripheral arterial occlusive disease (defined as either amputation or peripheral artery operation/stenting), myocardial infarction, bypass operation, or percutaneous coronary intervention.

**Supplementary Methods 3: Genotyping**

Genotyping and respective data cleaning was previously described in more detail.[6] In brief, DNA of 5 123 GCKD participants was isolated from whole blood and genotyped at 2 612 357 variants using the Illumina HumanOmni2.5 Exome BeadChip array (Illumina, GenomeStudio, Genotyping Module Version 1.9.4) at the Helmholtz Center Munich. In total, 89 samples were excluded based on QC steps regarding call rate, sex, heterozygosity, genetic ancestry and relatedness. On the variant level, single nucleotide polymorphisms (SNPs) were removed if either the call rate was <0.96, when the assumption of the Hardy-Weinberg equilibrium was violated (p-value$<1\times10^{-5}$), or when they were on duplicated positions.

Genotypes were imputed at the Michigan Imputation Server[7] (minimac3 v2.0.1) with the Haplotype Reference Consortium (HRC) haplotype version r1.1 as the reference panel, and Eagle 2.3 was used for phasing. The final genotype dataset contained 5,034 participants with information on 7 750 367 high-quality autosomal bi-allelic variants.

**Supplementary Methods 4: Polygenic Score Development and Validation**

We developed and tuned a PGS for log(eGFR) following a workflow published previously.[8] We randomly split 321 589 unrelated UK Biobank (application number:

17712) individuals of European ancestry into two groups, one containing 90% of the individuals that was used to conduct GWAS of log(eGFR) ($N_{GWAS}$ = 289 432), and one containing the remaining 10% of individuals that was used to select tuning parameters and validate the trained PGS models ($N_{validation}$ = 32 157).

We first ran a GWAS of log(eGFR) among the 289 432 UK Biobank participants, adjusting for age, sex, and the top 40 genetic principal components. We then used METAL to perform a meta-analysis to combine the UKB GWAS summary statistics with corresponding summary statistics from the CKDGEN Consortium[9], from which the ARIC and GCKD studies had been excluded to obtain a non-overlapping sample. From the results of the meta-analysis, approximately 1.5 million SNPs present on the Illumina Multi-Ethnic Genotyping Array (MEGA) Beadchip and HapMap3 were retained for score construction.[8]

The PGS was calculated using the LDpred algorithm.[10] Specifically, we created seven candidate LDpred PGSs corresponding to seven different pre-specified proportions of causal variants, $\rho$. This Bayesian approach utilizes GWAS summary statistics to compute the posterior mean effect sizes for the genetic variants by assuming a prior of the joint effect sizes and incorporating the LD structure calculated based on an external reference panel. In our case, the genetic data of 498 unrelated individuals in the 1000 Genomes Project was used as the LD reference.[11] With respect to user-specified parameters in LDpred, we used the default of 500 (the total number of SNPs divided by 3 000) for the LD radius, which is the number of variants being adjusted for at each side of a variant.[10] The fraction of causal variants, $\rho$, which can be selected via parameter tuning on a separate validation dataset, was tested at 1, 0.3, 0.1, 0.03, 0.01, 0.003, and 0.001, as suggested in Vilhjálmsson et al.[10]

For PGS tuning, the seven candidate LDpred PGS were calculated for the 32 157 independent individuals in the UKB validation dataset in order to select the best performing PGS. The best performing PGS along with the corresponding “optimal” value of ρ was selected based on $R^2$, i.e., the proportion of the variance of log(eGFR) explained by the PGS. Specifically, we fitted a linear regression model with log(eGFR) being the outcome, each candidate PGS being the exposure, and age at baseline, sex, and the first 40 PCs of genetic ancestry as the covariates.

**Supplementary Methods 5: PGS calculation in GCKD**

The eGFR PGS was calculated for GCKD participants with available genetic data using LDpred provided by Vilhjálmsson et al. (version 1.0.6, score option).[10] Subsequently, it was rescaled so that a higher eGFR PGS reflects lower eGFR to reflect that lower eGFR is harmful. Additionally, the eGFR PGS was standardised to a mean of 0 and a standard deviation of 1.

**Supplementary Methods 6: Statistical Analyses**

Cox regression models were fitted to evaluate the association of the eGFR PGS with the three main outcomes. All analyses were conducted for the eGFR PGS as a continuous variable, as well as for categories of the eGFR PGS, namely quartiles and deciles. The first 10 genetic principal components (PCs) were evaluated for potential inclusion as covariates and incorporated when they were nominally significantly ($p<0.05$) associated in a linear regression model with the eGFR PGS being the dependent variable and additionally adjusted for age and sex. Besides unadjusted analyses, models were adjusted in three incremental ways: 1) age + sex + significantly associated PCs, 2) model 1 + baseline eGFR and 3) model 2 + log(UACR).

Cox regression models provide results in the form of hazard ratio (HR) estimates for death, and cause-specific HR estimates for the other endpoints (KF, 3P-MACE) in the presence of the competing event (i.e., any death of other cause). The statistical significance threshold was set to p=0.05/3 to account for the three adjustments in the main outcome KF. In the competing event scenario for KF and 3P-MACE, subdistribution hazard analyses were carried out for comparison to detect possible indirect effects and obtain a summary measure of effect.[12,13] Graphical assessment of the proportional hazard assumption based on Schoenfeld residuals for the eGFR PGS showed no evidence for major violations.

Furthermore, we assessed whether the eGFR PGS carried predictive ability for the renal endpoint KF. The added predictive ability of the eGFR PGS was investigated in addition to the well-established 4-variable kidney failure risk equation (KFRE)[14,15] that is based on age, sex, eGFR and UACR. First, we investigated if the eGFR PGS added to model performance via a likelihood ratio test (LRT). Next, we compared the discriminative ability of the two nested models at years six of follow-up using the inverse probability censoring weighted c-index[16], a time-to-event equivalent to the area under the receiver operating characteristic (ROC) curve (AUC) value. For illustrative purposes, we plotted calibration plots showing predicted and observed estimated risks in deciles of the eGFR PGS at year six of follow-up, ROC curves at year six and prediction error curves over the six years of follow-up and integrated this curve to obtain the summary measure integrated prediction error curve (IPEC).[17,18] For the sake of more accurate performance measures, we provide 0.632+ estimates using 100 bootstrap samples for all prediction performance measures.[19]

### References

1. Eckardt KU, Bärthlein B, Baid-Agrawal S, et al. The German Chronic Kidney Disease (GCKD) study: design and methods. *Nephrol Dial Transplant Off Publ Eur Dial Transpl Assoc - Eur Ren Assoc*. 2012;27(4):1454-1460. doi:10.1093/ndt/gfr456

2. Titze S, Schmid M, Köttgen A, et al. Disease burden and risk profile in referred patients with moderate chronic kidney disease: composition of the German Chronic Kidney Disease (GCKD) cohort. *Nephrol Dial Transplant Off Publ Eur Dial Transpl Assoc - Eur Ren Assoc*. 2015;30(3):441-451. doi:10.1093/ndt/gfu294

3. Bycroft C, Freeman C, Petkova D, et al. The UK Biobank resource with deep phenotyping and genomic data. *Nature*. 2018;562(7726):203-209. doi:10.1038/s41586-018-0579-z

4. Köttgen A, Pattaro C. The CKDGen Consortium: ten years of insights into the genetic basis of kidney function. *Kidney Int*. 2020;97(2):236-242. doi:10.1016/j.kint.2019.10.027

5. Levey AS, Stevens LA, Schmid CH, et al. A new equation to estimate glomerular filtration rate. *Ann Intern Med*. 2009;150(9):604-612. doi:10.7326/0003-4819-150-9-200905050-00006

6. Li Y, Sekula P, Wuttke M, et al. Genome-Wide Association Studies of Metabolites in Patients with CKD Identify Multiple Loci and Illuminate Tubular Transport Mechanisms. *J Am Soc Nephrol JASN*. 2018;29(5):1513-1524. doi:10.1681/ASN.2017101099

7. Das S, Forer L, Schönherr S, et al. Next-generation genotype imputation service and methods. *Nat Genet*. 2016;48(10):1284-1287. doi:10.1038/ng.3656

8. Yu Z, Jin J, Tin A, et al. Polygenic Risk Scores for Kidney Function and Their Associations with Circulating Proteome, and Incident Kidney Diseases. *J Am Soc Nephrol JASN*. Published online September 21, 2021:ASN.2020111599. doi:10.1681/ASN.2020111599

9. Wuttke M, Li Y, Li M, et al. A catalog of genetic loci associated with kidney function from analyses of a million individuals. *Nat Genet*. 2019;51(6):957-972. doi:10.1038/s41588-019-0407-x

10. Vilhjálmsson BJ, Yang J, Finucane HK, et al. Modeling Linkage Disequilibrium Increases Accuracy of Polygenic Risk Scores. *Am J Hum Genet*. 2015;97(4):576-592.

doi:10.1016/j.ajhg.2015.09.001

11. Siva N. 1000 Genomes project. *Nat Biotechnol*. 2008;26(3):256. doi:10.1038/nbt0308-256b

12. Fine JP, Gray RJ. A Proportional Hazards Model for the Subdistribution of a Competing Risk. *J Am Stat Assoc*. 1999;94(446):496-509. doi:10.1080/01621459.1999.10474144

13. Latouche A, Allignol A, Beyersmann J, Labopin M, Fine JP. A competing risks analysis should report results on all cause-specific hazards and cumulative incidence functions. *J Clin Epidemiol*. 2013;66(6):648-653. doi:10.1016/j.jclinepi.2012.09.017

14. Tangri N. A Predictive Model for Progression of Chronic Kidney Disease to Kidney Failure. *JAMA*. 2011;305(15):1553. doi:10.1001/jama.2011.451

15. Tangri N, Grams ME, Levey AS, et al. Multinational Assessment of Accuracy of Equations for Predicting Risk of Kidney Failure: A Meta-analysis. *JAMA*. 2016;315(2):164. doi:10.1001/jama.2015.18202

16. Gerds TA, Kattan MW, Schumacher M, Yu C. Estimating a time-dependent concordance index for survival prediction models with covariate dependent censoring. *Stat Med*. 2013;32(13):2173-2184. doi:10.1002/sim.5681

17. Porzelius C, Binder H, Schumacher M. Parallelized prediction error estimation for evaluation of high-dimensional models. *Bioinforma Oxf Engl*. 2009;25(6):827-829. doi:10.1093/bioinformatics/btp062

18. Binder H, Schumacher M. Adapting prediction error estimates for biased complexity selection in high-dimensional bootstrap samples. *Stat Appl Genet Mol Biol*. 2008;7(1):Article12. doi:10.2202/1544-6115.1346

19. Efron B, Tibshirani R. Improvements on Cross-Validation: The .632+ Bootstrap Method. *J Am Stat Assoc*. 1997;92(438):548. doi:10.2307/2965703
